# Respiration recording for fMRI: breathing belt versus spine coil sensor

**DOI:** 10.1101/2023.08.24.23294529

**Authors:** Marilena Wilding, Anja Ischebeck, Natalia Zaretskaya

**Affiliations:** Department of Psychology, University of Graz, Universitätsplatz 2, 8010 Graz, Austria; BioTechMed-Graz, Mozartgasse 12, 8010 Graz, Austria

## Abstract

Physiological signals such as pulse and respiration strongly contribute to non-neuronal signal change of the blood oxygenation level-dependent (BOLD) contrast in functional magnetic resonance imaging (fMRI). This has been observed not only during task-based but also during resting-state fMRI measurements, where the confounding influence of physiological signals is most pronounced. Over the last decades, a variety of techniques evolved, aiming at detecting and removing physiological artifacts in fMRI time series. These follow either a solely data-driven approach or rely on externally recorded physiological data. To record cardiac and respiratory signals, typically pulse oximetry or electrocardiography (ECG) and a respiration belt are used, respectively. New technologies allow to capture respiratory signal directly with a sensor placed within the spine coil in the patient table, eliminating the need of a respiration belt, which considerably increases participants’ comfort. However, little is known about the effectiveness of these new technologies and how they compare to the standard respiration belt recording. In the current study, we compared the two devices, respiration belt and spine coil sensor, in their suitability for physiological noise removal during a visual perception task and during rest. We did not find any differences in resting-state functional connectivity (RSFC) or stimulus-related activity between data corrected with the two recording devices. However, we did find reduced residual noise in the time series corrected with spine coil-derived respiration signals compared to belt-based corrected data in the task dataset. Our results show that spine coil-derived respiration recordings are slightly superior to belt respiration recordings for physiological noise removal in task-induced activity, with spine coil recordings having an additional advantage in terms of subject comfort.

## 1. Introduction

The blood oxygenation level-dependent (BOLD) contrast is one of the most frequently used indicators of brain activity in human neuroimaging. Although it is an indirect measure of neuronal events, it nevertheless reflects changes in the oxygenation of the blood that are evoked by neuronal activity. This signal, however, doesn’t only indicate neuronal activity. It further contains several confounding noise components that obscure the main signal. Multiple noise sources have been identified over the last decades, such as background noise, thermal noise (system-related noise), instrumental drifts, instabilities of hardware, participant’s head motion as well as physiological signals, such as cardiac rate, respiratory rate and resultant changes in the level of CO_2_ (Caballero-Gaudes and Reynolds, 2017; Keilholz et al., 2017; Liu, 2016).

Participant motion and physiological noise have been found to contribute most to non-neuronal signal change in the BOLD contrast (Caballero-Gaudes and Reynolds, 2017; Kasper et al., 2017; Keilholz et al., 2017). Studies suggest its contribution to the signal variance to be comparable to that of the BOLD signal fluctuations during a task and at rest (Caballero-Gaudes and Reynolds, 2017). Task-based fMRI is thought to be more robust to non-neuronal noise since the observed effects are usually a result of averaging over multiple trials. However, this view fails to consider the notion that changes in cardiac rate or breathing rate can correlate with the examined task, especially when it is emotionally arousing or cognitively challenging, and hence introduce additional noise in a task-locked fashion (Birn et al., 2009; Bright et al., 2014a). In resting-state functional connectivity (RSFC) analysis, the influence of physiological noise is even more pronounced, as non-neuronal noise has been found to introduce spurious correlations between voxels’ time series (Birn, 2012; Murphy et al., 2013), which can even mimic functional brain networks (Bright et al., 2020; Chen et al., 2020). Although physiological noise is typically observed at frequencies higher than the usual BOLD signal fluctuations at rest (i.e., cardiac: ∼1 Hz, respiratory: ∼0.3 Hz, BOLD: < 0.1 Hz), due to aliasing effects present at repetition times (TRs) of ≥ 2 s, the physiological signals can still appear in the low-frequency segment of the power spectrum, where they are hard to distinguish from low-frequency oscillations of neural origin (Bhattacharyya and Lowe, 2004; Murphy et al., 2013).

### Effects of cardiac and respiratory noise

Cardiac pulsation activity is thought to have a substantial effect on the relative distribution of fluid components (e.g., cerebrospinal fluid, blood) and also more solid brain tissue within the skull (Bhattacharyya and Lowe, 2004; Dagli et al., 1999). Expectedly, its effects are most noticeable near major blood vessels, as an increase in blood pressure intensifies pulsation of the vessels and hence causes local movements (Dagli et al., 1999; Glover et al., 2000). Changes in the cardiac rate were also found to be related to variations in the BOLD signal amplitude, which is not surprising since both signals are dependent on cerebral blood flow (CBF), cerebral blood volume (CBV), and oxygenation. This close relationship, however, can introduce confounds not only around large vessels, but also within the gray matter tissue (Chang et al., 2009; Shmueli et al., 2007).

Respiratory activity can affect the BOLD signal in various ways. First, abdominal motion can lead to perturbation of the magnetic field (by inducing changes in field strength and homogeneity), which can cause distortion of the acquired data (Raj et al., 2001). Small head movements during breathing can further produce spin history effects, that can last for several volumes and are spatially dependent on the axis of the movement (Caballero-Gaudes and Reynolds, 2017; Friston et al., 1996; Muresan et al., 2005). In addition to effects related to motion, even small and ordinary variations in respiratory volume and rate can induce considerable changes in the low-frequency BOLD signal. This is due to the fact that changes in breathing are accompanied by shifts in the arterial level of carbon dioxide (CO_2_), which, as an important vasodilator, modulates local cerebral blood flow (Birn et al., 2006; Wise et al., 2004). This is especially relevant near locations with high blood volume, such as blood vessels and gray matter (Birn et al., 2008). Together, these findings highlight the complexity of distinguishing signal driven by neural activity from mere physiological noise, which can affect the BOLD signal in a broad variety of ways.

### Minimizing physiological noise

Multiple techniques to determine and, most importantly, remove physiologically induced noise in BOLD time series have evolved over the last years (Agrawal et al., 2020; Caballero-Gaudes and Reynolds, 2017; Murphy et al., 2013), mainly following one of the following approaches or their combination. On the one hand, data-driven models aim to approximate the contribution of physiological fluctuations based on signals from regions which are not likely to be effectively influenced by neuronal activity, such as cerebrospinal fluid (CSF) and white matter. These estimations, however, only allow to remove the average signal of said regions, approximating physiological contribution to gray matter signal and fitting it to every gray matter voxel. Based on this principle, more advanced methods such as CompCor (component based noise correction method) have been developed, that enable modelling the physiological signal in gray matter as multiple principle components of the CSF and white matter time courses instead of using the average signal (Behzadi et al., 2007).

Reference-based approaches, on the other hand, aim at estimating the physiological noise not from indirect sources but from data acquired using external measurement devices (such as pulse oximeter, respiration belt, or electrocardiogram), which directly record physiological signals throughout the fMRI session. These signals are then retrospectively synchronized with the functional data (Kasper et al., 2017) in order to estimate the influence of physiology on each voxel’s time series. One widely established example is the RETROICOR (retrospective image correction) algorithm, which aims to determine the phase of each cardiac and respiratory cycle whenever a slice is acquired. Subsequently, the periodic effects of the physiological noise are modelled using a Fourier expansion of the phases that is fit to the time series of each voxel (Glover et al., 2000).

Despite the substantial effects of respiration on the BOLD signal (Birn et al., 2008; Caballero-Gaudes and Reynolds, 2017; Chang et al., 2009; Liu, 2016; Murphy et al., 2013; Verstynen and Deshpande, 2011), little is known about the different means of physiological signal recording and their suitability for subsequent physiological signal modelling. Traditionally, respiratory activity is recorded using a respiration belt that is applied around an individual’s upper torso near the diaphragm and registers the shift in abdominal volume. The new BioMatrix technology implemented in more recent models of the Siemens MRI scanners, however, additionally allows the recording of respiration via a sensor embedded in the spine coil within the patient table (Runge et al., 2019). The sensor produces a local magnetic field that changes with respiration-induced motion. This option is fully integrated into the scanner’s structure and hence eliminates the necessity for respiration belt usage, which increases subjects’ comfort. However, it remains unclear whether the new technology reflects the respiratory signal in a comparable manner and is equally suitable for modelling physiological artifacts in fMRI experiments. To investigate this, we simultaneously recorded the two types of signals in one task-based and one resting-state fMRI dataset, and formally compared them in their suitability for minimizing physiological noise. We could show that both recording methods are similarly suitable for physiological noise removal. Importantly, we did not find any differences between data corrected with both denoising methods, for resting-state functional connectivity as well as stimulus-related activity. Examination of residual noise, however, revealed less remaining noise in the data denoised with the *spine* coil sensor recording compared to the data denoised using signal from the respiratory *belt* in the task dataset.

## 2. Material and Methods

### 2.1 Participants

The task-based dataset comprised 25 participants between 19 to 31 years (12 female, mean age = 23.61, *SD* = 3.40) and the resting-state dataset included 56 volunteers between 19 and 35 years (35 female, mean age= 24.22, SD= 3.15) after excluding 6 and 4 participants, respectively, either due to insufficient physiological signal quality or technical issues during recording. Resting-state measurements were followed by several task runs, which are not discussed in this paper. Participants had no neurological, psychiatric, or cardiovascular diseases and were not taking any medication on a regular basis. Before data acquisition, they gave written informed consent and were instructed in a standardized manner. Both studies were conducted following the Declaration of Helsinki and were approved by the local ethics committee of the University of Graz. Detailed descriptions of the two datasets are provided in our previous studies (task: Wilding et al., 2022, rest: Wilding et al., 2023).

### 2.2 Data acquisition

#### fMRI data

fMRI data for both datasets were acquired on a 3T Siemens Magnetom Vida scanner (Siemens Healthiness, Erlangen, Germany) using a 64-channel head coil. Functional images were obtained using the blood oxygenation level dependent (BOLD) contrast. For detailed acquisition parameters, see Table 1.

#### Rest

The resting-state dataset was acquired in an 8-minute scanning session. Participants were instructed to fixate a red dot in the center of the screen and avoid any directed cognition.

#### Task

Task-related data were acquired using a multiband echo planar imaging sequence (EPI). During the experiment, participants were repeatedly presented with a bistable motion stimulus for a duration of 1 second, interleaved with long (25-50 s) baseline periods. Immediately after the stimulus presentation they had to report their spontaneously occurring percept via a button press. Each participant underwent six functional runs of around 13 minutes, the exact duration depending on the randomization of the inter-stimulus periods. This resulted in of a total measuring time of approximately 75 minutes.

**Table 1.** fMRI acquisition parameters for both datasets.

|  | *Rest* | *Task* |
| --- | --- | --- |
| TR | 3.22 s | 0.88 s |
| TE | 0.32 s | 0.30 s |
| multiband factor | - | 3 |
| acceleration factor | - | GRAPPA, 2 |
| voxel size | 3 x 3 x 3 mm | 3 x 3 x 3 mm |
| flip angle | 82° | 65° |
| FOV | 228 mm | 210 mm |
| number of slices | 46 | 45 |
| number of volumes | 155 | on avg. 5100 |

#### Physiological data

Physiological measures in both studies were recorded throughout the whole fMRI data acquisition period. Heart rate was recorded with a photoplethysmograph clip at the participants’ left index finger at a sampling rate of 400 Hz. This technique allows to monitor and record an individual’s pulse by measuring the absorption of infrared light in body tissue, which is modulated by the cardiac cycle (Nilsson, 2013).

The respiration signal was captured simultaneously by two devices, the respiration belt (*belt*) and the spine coil sensor (*spine*). The respiration belt is placed around the upper torso, and an air cushion placed between the belt and the torso that detects pressure changes induced by participants’ chest contraction and expansion (sampling rate: 400 Hz). The spine coil respiration sensor is embedded in the BioMatrix spine coil of the Siemens Magnetom Vida scanner (Runge et al., 2019). It applies a local electromagnetic field near the torso, which differs from the Larmor frequency (i.e., 30 MHz) and hence does not interfere with the main scanner signal. This electromagnetic field varies with motion produced by the different phases of the respiratory cycle. Surrounding coil elements detect these changes, which reflect the participant’s breathing pattern. The recording of physiological signals was performed using the command line tool *ideacmdtool* by logging pulse and respiration signals throughout the fMRI data acquisition. This resulted in two files, one containing pulse, and one containing respiration data, which were used for further processing.

### 2.3 Data analysis

#### 2.3.1 Processing of the physiological data

Physiology regressors were obtained from the physiological log-files using the MATLAB-based *PhysIO* toolbox (Kasper et al., 2017). In the first step, respiratory data quality was checked by inspecting raw data (Fig. 1A) as well as the distribution of breathing amplitudes, allowing us to examine signal quality and detect potential issues with signal recording. Sufficient respiratory signal quality is reflected in a high number of low/intermediate amplitudes with a long tail towards higher amplitudes, which correspond to sporadic deep breaths (Fig. 1B). In the case of technical problems during recording, such as ceiling effects in the respiratory signal (e.g., belt was too tight) or prolonged signal loss (e.g., temporary detachment of belt, see Fig. 1C), there is a distribution peak at the maximum amplitude or around zero, respectively. Cardiac signal was examined for possible periods of detachment by visually inspecting the data. Next, the cardiac and respiratory signal was aligned with the fMRI time series using the system time stamps and an algorithm performing iterative peak detection was applied to identify repetitive signal features and discard compromised segments and noise. Problematic time series were excluded when artifactual signal segments were present in ≥ 40 % of the signal, which was the case in three participants of each dataset. The remaining datasets were processed with RETROICOR phase expansion algorithm (Glover et al., 2000), which allows to consider variation within the physiological signals by modelling the periodic effects of pulsatile motion and field fluctuations as Fourier expansion of the respiratory phase. The expansion orders were set following the parameters of Harvey et al. (2008), i.e.,3^rd^ order cardiac model, 4^th^ order respiratory model, and 1^st^ order interaction model. This resulted in 6 cardiac phase regressors, 8 respiratory phase regressors, and 4 interaction terms; in total 18 regressors per device (RETROICOR*_belt_* and RETROICOR*_spine_*). Finally, regressors were downsampled to a respective acquisition TR and added to the GLM.

Signal correspondence of the unprocessed respiratory signal recorded using *belt* and *spine* was quantified using Spearman’s rank correlation in MATLAB R2019b (MathWorks, Inc., Natick, MA).

**Figure 1.**
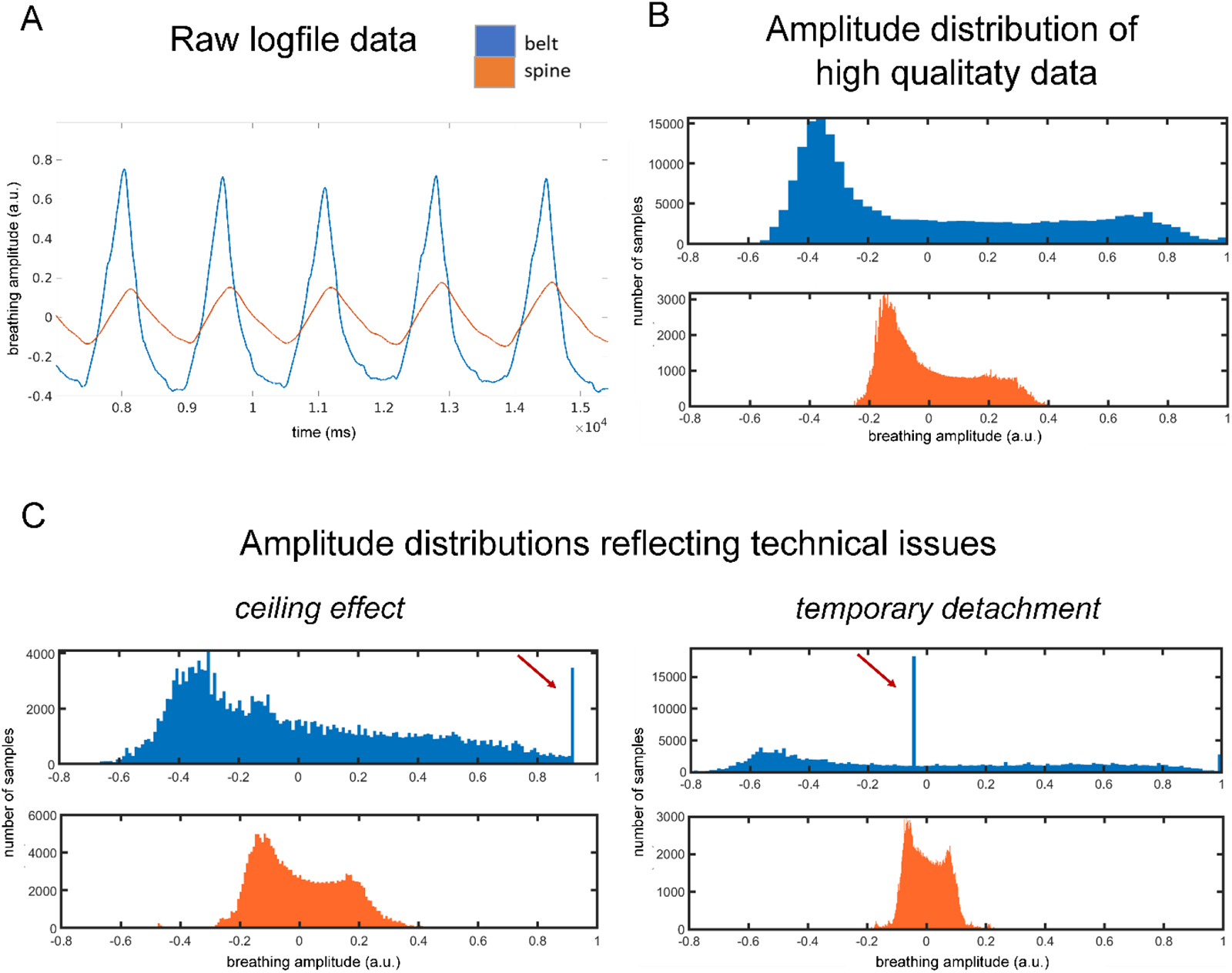
Comparison of respiratory signal captured with a breathing belt (*belt*; blue) or spine coil sensor (*spine*; orange). A) Example raw signal for both recordings showing five breathing cycles. B) Typical distribution of amplitudes in the raw signal for both recordings, both following a right-skewed distribution. C) Example of typical problems during the respiratory signal recording (indicated by red arrows) reflecting ceiling effects (belt was too tight) and temporary detachment of the belt, which are visible only in the *belt* recording.

#### 2.3.2 Preprocessing of fMRI data

Structural and functional data of both datasets were preprocessed using fMRIPrep version 20.2.3 (Esteban et al., 2019), which is based on Nipype 1.6.1 (Gorgolewski et al., 2011). The description of preprocessing steps is based on the fMRIPrep boilerplate text.

##### Anatomical data

The T1-weighted image was corrected for intensity non-uniformity and used as T1w-reference throughout the following steps. Next, the T1w-reference was skull-stripped and brain tissue segmentation into cerebrospinal fluid (CSF), white matter (WM) and gray matter (GM) was performed on the brain-extracted T1-weighted image.

##### Functional data

First, a reference volume and its skull-stripped version were generated. Next, a B0-nonuniformity map was estimated based on echo-planar imaging (EPI) references with opposing phase-encoding directions. Based on the estimated susceptibility distortion, a corrected EPI reference was calculated for a more accurate co-registration with the anatomical reference. The BOLD reference was co-registered to the T1w reference by applying boundary-based registration (*bbregister;* (Greve and Fischl, 2009) with 6 degrees of freedom. Head-motion parameters with respect to the BOLD reference (transformation matrices for the six rotation and translation parameters) were estimated before any spatiotemporal filtering using *mcflirt* (FSL 5.0.9). The BOLD time-series were resampled onto MNI space by applying a single, composite transform to correct for head-motion, slice acquisition timing and susceptibility distortions.

The preprocessed datasets were then imported into the CONN toolbox (Whitfield-Gabrieli and Nieto-Castanon, 2012); version CONN21.a), which uses SPM 12 (http://fil.ion.ucl.ac.uk/spm/), and analyzed separately.

#### 2.3.3 Resting-state data analysis

Resting-state data analysis was performed using CONN version CONN21.a), which uses SPM 12. Functional data were smoothed using spatial convolution with a Gaussian kernel of 6 mm full width half at maximum (FWHM). Potential outlier scans were identified using artifact detection software (ART) as volumes with framewise displacement above 0.9 mm or global BOLD signal changes above 5 standard deviations. Next, the functional time series were denoised using a standard denoising pipeline (Nieto-Castanon, 2020) including the regression of potential confounding effects characterized by 6 motion parameters (derived from fMRIPrep) and their first order derivatives (6), outlier scans (15 regressors or less, depending on the participant), and physiology regressors derived from the RETROICOR model (18). This was followed by bandpass filtering of the BOLD timeseries between 0.008 Hz and 0.09 Hz. To compare the effect of the respiration recording method on the resting-state data, functional runs of each subject were added as three separate sessions that only differed in the type of physiology regressors (RETROICOR*_belt_,* RETROICOR*_spine_* or *none*).

##### Functional connectivity analysis

To characterize the patterns of functional connectivity, ROI-to-ROI connectivity matrices (RRC) were estimated using the 164 HPC-ICA network parcellation, which is implemented in CONN. These matrices reflect the strength of functional connectivity between each pair of ROIs. Functional connectivity strength was represented by bivariate Fischer-transformed Pearson’s correlation coefficients, defined separately for each pair of target areas. First, we compared resting-state functional connectivity from data that was not corrected for physiological noise to data corrected with either *belt* or *spine*. In the next step, we directly compared functional connections in data corrected with *belt* or *spine*. Results were thresholded using a connection-level threshold of p < 0.01, and a cluster-level FDR-corrected threshold of p < 0.05.

##### Residual noise analysis in the resting-state data

To examine differences in remaining noise after physiology correction with either recording, we compared the standard deviation of residuals between both. The denoised signal files were automatically created by the CONN toolbox after denoising and the standard deviation of residual voxel time course over time was calculated for each 4-D volume using an *fslmaths* command in FSL version 6.0.4 (Smith et al., 2004). To evaluate the general effects of physiological noise modelling compared to no physiology correction, we performed paired t-tests between *none* and *belt/spine* using the respective standard deviation maps in SPM 12. The results were corrected for multiple comparisons using a threshold of p < 0.01, FWE corrected. To directly compare residual noise in the *belt*- and *spine*-corrected data, a voxel-wise paired t-test was calculated between *belt* and *spine* datasets. Since the effects of direct comparison are expected to be more subtle, statistical maps were thresholded using a more liberal voxel-level threshold of p < 0.01, uncorrected.

#### 2.3.4 Task data analysis

Task-based data analysis was performed using the CONN toolbox (version 21.a), which uses SPM 12. The task-based functional data were smoothed using spatial convolution with a Gaussian kernel of 8 mm full width at half maximum. The first four volumes of each run were discarded to allow for T1 equilibration effects.

##### Activity analysis

For the first-level analysis, stimulus onsets were modelled as events of 2 s duration as regressors of interest. In addition, motion parameters (6) and physiology regressors (18) were included into the GLM as nuisance regressors. The first-level GLM analysis was conducted twice, once with RETROICOR_belt_ and once with RETROICOR_spine_ nuisance regressors. Beta values for the stimulus onset of each GLM and each participant were used for the subsequent second-level group analysis. First, we examined stimulus-related activity in both models separately. To test whether there is an overall activity difference between *belt* and *spine*, a whole-brain analysis using the t-contrast “*belt* – *spine”* was performed on the second level. Results were thresholded using a voxel-level threshold of p< 0.01 and a cluster-level FDR-corrected threshold of 0.05.

##### Standard deviation of residuals analysis

To confirm our findings for the resting-state fMRI, we performed a similar analysis of residuals for the task-based fMRI dataset. To account for fluctuations related to task, in addition to including the noise regressors we also included the effects of task in the GLM before examining the standard deviation of the residuals. To directly compare *belt* and *spine*, we performed a voxel-wise paired t-test on the standard deviation maps using a voxel-level threshold of p < 0.01, uncorrected.

All figures depicting functional results were created using the MATLAB-based *bspmview (v.20180918)* program (Stund, 2016).

## 3. Results

### 3.1. Correspondence of signals

First, we examined the correspondence between the raw respiratory signal acquired with *belt* and *spine* for the resting-state dataset. A Spearman’s rank correlation revealed a high correspondence between the two signals, with a median correlation of R= 0.69 between both signals over all participants (min.: R= 0.36, max. R= 0.89; IQR= 0.11), see Figure 2.

**Figure 2.**
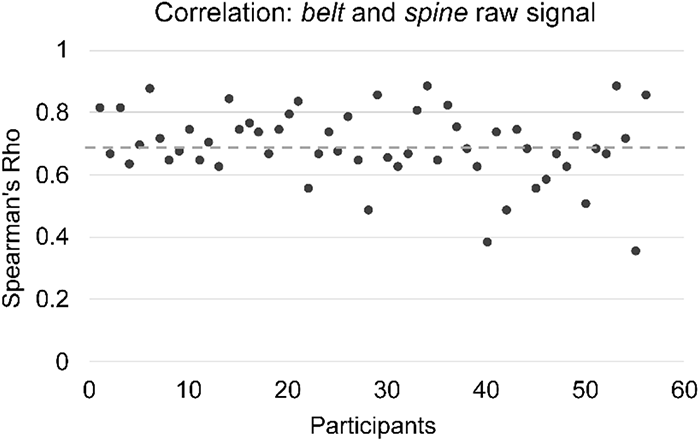
Correlation between the respiratory signal recorded with *belt* and *spine* devices. The median correlation across participants (R= 0.69) is depicted by the grey dotted line.

### 3.2. Effects of noise correction

#### 3.2.1 Functional connectivity

We examined the general effect of physiological noise modelling on functional connectivity by comparing functional connectivity derived from data denoised without physiology regressors (*none*) with functional connectivity corrected with either RETROICOR*_belt_* or RETROICOR*_spine_*. The comparison “*none* – *belt”* revealed significantly higher cortical ROI-to-ROI functional connectivity in the absence of physiological modelling, suggesting the presence of artifactual correlations produced by physiological noise (Fig. 3A). Similar results were found for the comparison “*none* – *spine”*. For the latter, we found a slightly higher number of significant functional connections than for “*none* – *belt”*. We then directly compared resting-state functional connectivity derived from data corrected with either RETROICOR*_belt_* or RETROICOR*_spine_*. Our ROI-to-ROI functional connectivity analysis did not reveal any significant differences (even at a more liberal threshold of p < 0.01, uncorrected).

#### 3.2.2 Functional activity

For the task-dataset, we examined stimulus-related activity for data corrected with signal from each type of device. Our GLM analysis revealed a highly similar activity pattern in response to the stimulus for models containing RETROICOR*_belt_* or RETROICOR*_spine_* regressors. As illustrated in Figure 3B, we observed a widespread stimulus-related activity increase in areas overlapping the fronto-parietal network and the salience network, and an activity decrease in regions that are similar in location to the nodes of the default mode network.

Next, we investigated differences in stimulus-related activity between both denoising types. We did not observe any significant differences in stimulus-related activity between *belt* and *spine*, even at a liberal threshold of p<0.01, uncorrected (Fig. 3B).

**Figure 3.**
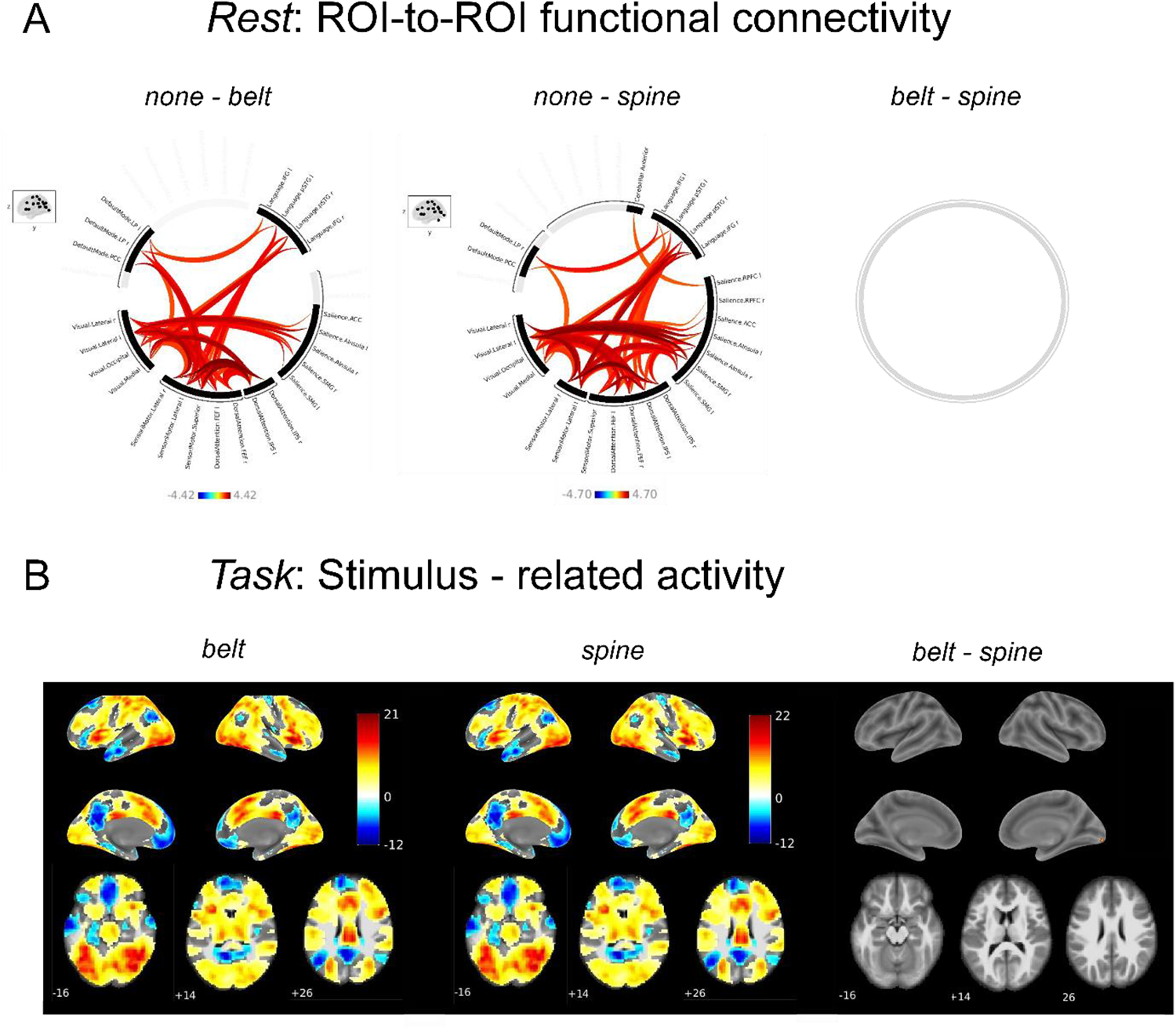
Effects of noise correction for both datasets. A) ROI-to-ROI functional connectivity differences between A) *none* and *belt*, and b) *none* and *spine*. Connection threshold: p < 0.01, cluster threshold: p < 0.05, FDR-corrected. B) Stimulus-evoked activity for both types of denoising; voxel-threshold: p< 0.01, cluster-threshold: p<0.05, FDR-corrected.

### 3.3 Residual noise

#### 3.3.1 Residual noise *resting-state*

We first confirmed that physiological noise modelling indeed reduces physiological noise in the data. To compare the amount of remaining noise after physiology correction (RETROICOR*_belt_* or RETROICOR*_spine_*) with no physiology correction (*none*), the temporal standard deviation of the denoised signal which included physiology regressors was compared with the standard deviation of the denoised signal that did not include physiology regressors. As expected, residual noise was higher without physiological noise modelling. There was a statistically significant difference between *none* and *belt* as well as *none* and *spine* throughout the whole brain volume, with higher standard deviation values for denoising without physiology regressors (both T_55_ 6.18, p<0.01, FWE corrected; see Figure S1).

To examine which device led to a more effective physiological noise removal in the resting-state data, we directly compared the standard deviation of residuals for the *belt*-denoised and *spine*-denoised data using a paired t-test. We found no significant differences between the standard deviation of the denoised signal of RETROICOR*_belt_* and RETROICOR*_spine_* at a threshold of p < 0.01, FWE-corrected. At an uncorrected threshold of p <0.01, our paired t-test revealed several positive (16, average voxel size= 43) and negative (12, average voxel size= 32) clusters for the contrast “*belt* – *spine”* (T_55_>= 2. 396; see Figure 4A).

#### 3.3.2 Residual noise *task*

Finally, we examined possible differences in the remaining noise for both denoising types in the task dataset, finding no significant differences. At the uncorrected level (p <0.01) our paired t-test revealed a clear distribution towards positive values (T_24_= 2.49; see Figure 4B), indicating more remaining noise in the *belt*-denoised data.

**Figure 4.**
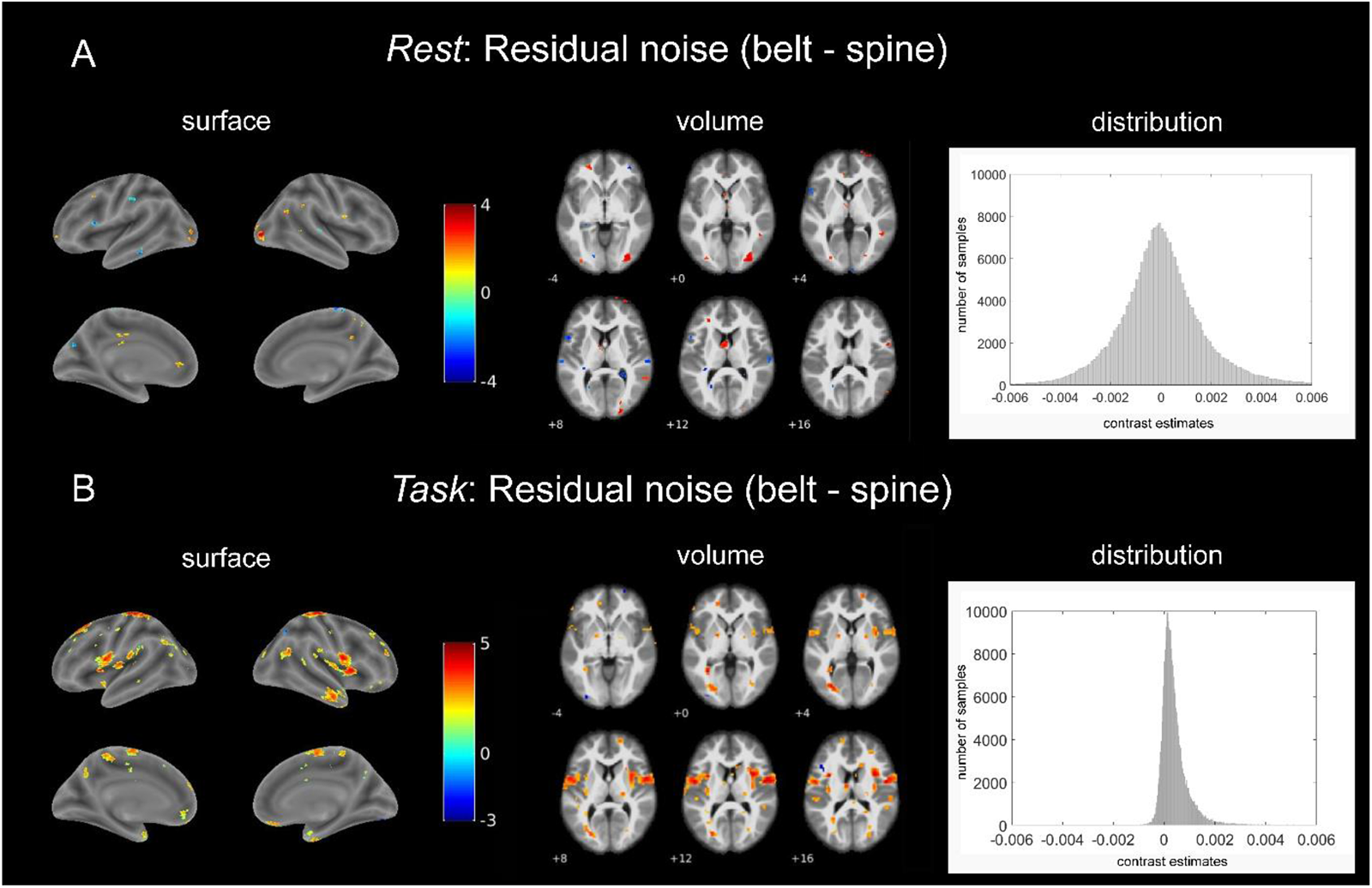
Comparison of residual noise in *belt*-denoised and *spine*-denoised data in the resting-state dataset (A) and in the task-based dataset (B). We observed higher noise values in the *belt*-denoised data, mainly in the task-dataset. Insets on the right show the distribution of contrast estimates for the contrast “belt vs. spine” within the brain mask for each dataset. The rest dataset resembles a normal distribution while in the task dataset a clear shift to positive values is visible.

## 4. Discussion

### Summary

Physiological noise is one of the main sources of non-neuronal noise in the BOLD signal. Recording physiological signals such as pulse and respiration during fMRI acquisition can minimize their impact on functional data by removing the main components of noise retrospectively. In this article, we compared two different respiratory signal recording devices for their overall ability to reduce physiological noise in fMRI data. To evaluate the consistency of the effects, we assessed their impact in a resting-state dataset and a task-based dataset. Our results show that the new spine coil sensor technology is as good as the conventional respiratory belt recording as a source signal for physiology denoising, and potentially even superior for the task-based dataset.

### Results

Our results emphasize the importance of including physiological noise correction in fMRI analyses, as they showed significantly more residual noise in data without any physiological correction compared to data denoised with either RETROICOR*_belt_* or RETROICOR*_spine_*. This effect was consistent for denoising using signals from both devices and was present throughout the brain. At the same time, our results suggest a potential advantage of the spine coil sensor for fMRI signal denoising. Specifically,denoising with RETROICOR_spine_ led to smaller variance in denoised task-induced signal compared to *belt*, although there was no clear trend in the resting-state data. Consistently, a slight trend towards superior *spine*-derived denoising was observable in the functional connectivity results. When comparing no correction (*none*) with *belt-* and *spine-*derived denoising, we found that data corrected with RETROICOR*_spine_* revealed fewer connected network nodes compared to data denoised with RETROICOR_belt_. Since physiological noise is known to manifest itself as spurious correlations between voxels, correlation/connectivity reduction may indicate a more rigorous removal of physiological noise with RETROICOR*_spine_*. However, these results must be interpreted with caution, as a direct comparison of *belt* and *spine* functional connectivity and task-based activity did not reveal any significant differences, even at liberal statistical thresholds.

### Potential reasons for differences

Our results imply a slight advantage of the spine coil sensor in correcting for physiological noise in task-based data. Reasons for this apparent discrepancy in the effectiveness of noise correction may be found at the raw signal level. The respiration belt needs to be applied at the correct position on the subject’s torso with an optimal degree of tightness before recording. However, if the strap deviates too much from the target position or if it is applied too loosely or too tightly, signal artifacts are likely to occur. For example, a strap that is too tight can result in ceiling effects, and incorrect positioning (also due to subject motion) can cause belt detachment or the air cushion slipping out from underneath the belt and consequent loss of signal (see Fig. 1 C and D). In contrast, spine sensors are implemented in the scanner table which not only increases acquisition comfort but also eliminates artifacts due to incorrect arrangement of device components. Here, we excluded the *belt* signal in six participants that exhibited severe artifacts. However, less severe cases were still included in the study and may have had an overall impact on signal quality between devices. This is also reflected in the signal correspondence between both devices, which is high (median R= 0.69), but not perfect and shows some variation across participants. It is noteworthy that no participant was excluded because of the low quality of the *spine* coil sensor signal, which again speaks in favor of this technology.

But why were device-related effects mainly present in the task-induced dataset? As shown in Figure 4 (right), the distributions of contrast-specific weighted combinations of beta values for both datasets differ in their appearance. While the distribution of contrast estimates in the task-dataset showed a clear rightward shift, the resting-state data resembles a Gaussian distribution with a center around zero. We speculate, that the difference in the amount of data, which was clearly in favor of the task-dataset, may partly obscure similar effects in the resting-state dataset due to less statistical power. While the resting-state dataset contained only 8 minutes of data with 155 volumes, due to its experimental nature and multiband acceleration the task-based dataset contained 75 minutes and 5100 volumes on average.

Aside from that, we consider it possible that additional effects of physiology may have been time-locked to the stimulus or task, which can typically be the case in arousing or cognitive challenging tasks and introduce systemic confounds to the data. As shown in Wilding et al., 2022, arousal levels in the task-dataset clearly increased after stimulus onset, reflecting strong stimulus/task-related physiological modulations. Previous studies showed that accounting for these physiological modulations e.g., by adding regressors based on the recorded physiological signal, can drastically improve the detection of the real neural signal (Birn et al., 2009; Bright et al., 2014b; Hillenbrand et al., 2016; Lane et al., 2009). Based on these findings, we suggest that due to a more efficient correction of stimulus/task-correlated confounds, task-related data corrected with RETROICOR_spine_ may show less residual noise compared to belt-based data as well as resting-state data.

### Limitations and future implications

In this article, we show that functional noise correction with spine coil sensor data appears to perform better in terms of residual noise than the belt signal, but mainly in task-induced activity. In the current study, we can’t completely rule out that this discrepancy between dataset is due to the amount of data in both datasets, as discussed above. Therefore, future studies that include longer resting-state measurements with multiple runs are needed to confirm that the advantages of the spine coil sensor are not evident in the resting-state data.

Moreover, as it is common practice when correcting for physiological noise, we used the RETROICOR algorithm to create noise regressors that allow to account for periodic effects in physiological noise. Although being a highly recommended option, there are several other reference-based approaches that can be used for physiological noise correction, which focus on slightly different physiological parameters, e.g., the respiratory response function, which accounts for variations in cardiac and respiratory rate or the breathing depth and end-tidal CO2 changes (Birn et al., 2008, 2006b; Chang and Glover, 2009). Future studies could therefore implement different physiological noise models/parameters to support and complement present results. Moreover, additional devices that are becoming more common for respiratory recording, such as the pulse oximeter (Addison et al., 2012; Dehkordi et al., 2018), should be compared with the breathing belt and/or spine sensors with respect to their suitability for respiration recording.

### Conclusion

Our study confirms that both types of devices can be effectively used to reduce physiological noise in resting-state and task-based fMRI data. Moreover, it suggests that models derived from the spine coil sensor data perform slightly better in removing noise of physiological origin compared to a conventional breathing belt recording, especially in task-induced activity.

## Data Availability

All data produced are available online at https://osf.io/hr7v4/

## 5. Important stuff

### Data and code availability statement

Data and code are available on OSF at: https://osf.io/hr7v4/.

### Funding

This work was supported by the BioTechMed-Graz Young Research Group Grant to N.Z.

## Acknowledgements

We thank Erik Fink and Thomas Zussner for their support in data collection.

## CReDIT authorship contribution statement

**M. Wilding:** Conceptualization, Methodology, Software, Formal analysis, Investigation, Writing – original draft, Visualization.

**A. Ischebeck:** Writing – review & editing, Supervision.

**N. Zaretskaya**: Conceptualization, Methodology, Resources, Writing – review & editing, Supervision, Funding acquisition.

## Supplementary Material

**Figure S1.**
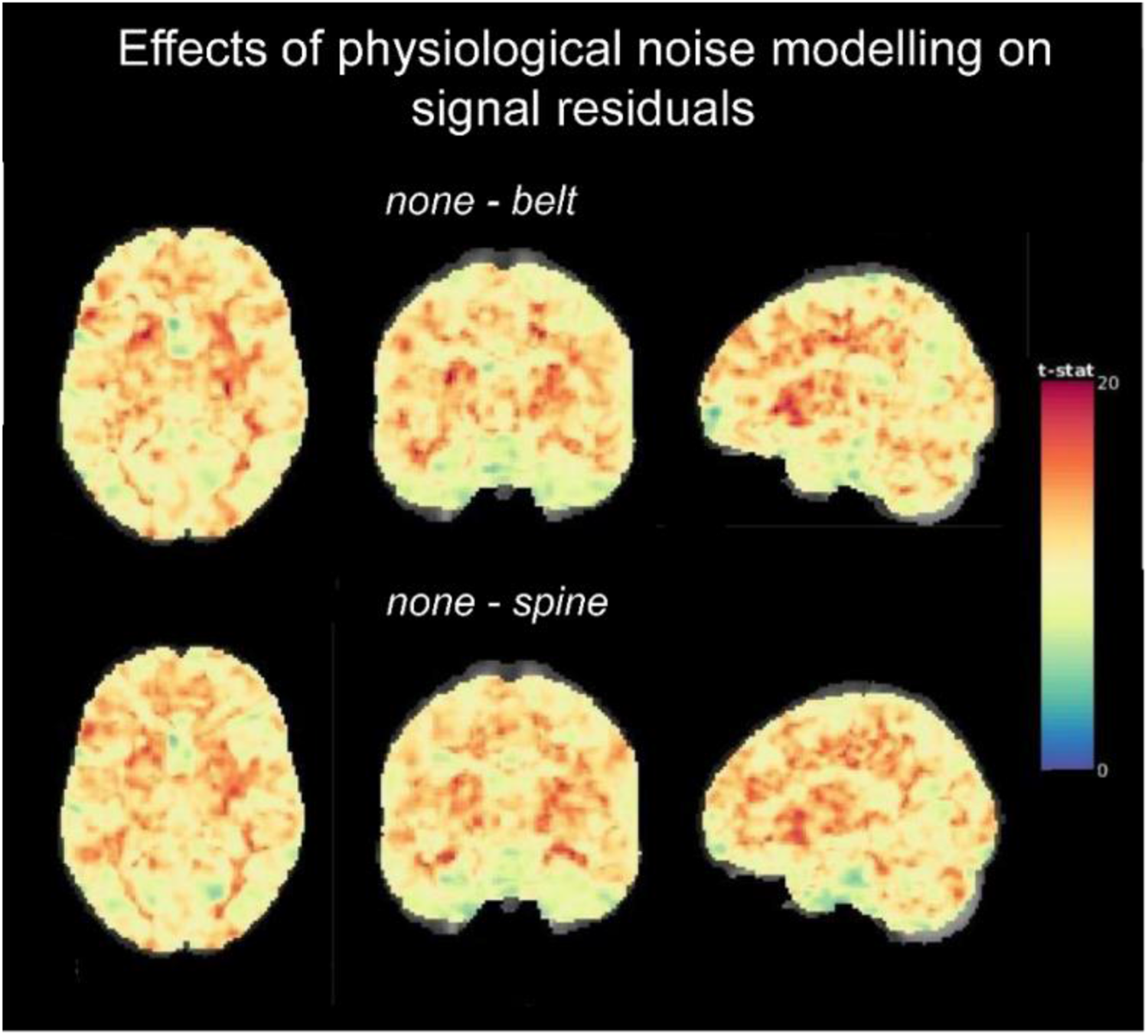
Standard deviation of model residuals for *none* versus *belt* (left) and *none* versus *spine* (right) in the rest dataset.

## Notes

### Competing Interest Statement

The authors have declared no competing interest.

### Funding Statement

This study was funded by the BioTechMed-Graz Young Research Group Grant to N.Z.

### Author Declarations

The local ethics committee of the University of Graz approved this work.

